# CBFB mutations predict endocrine therapy benefit in estrogen receptor–positive breast cancer

**DOI:** 10.64898/2026.05.18.26353467

**Authors:** Adar Yaacov, Gal Passi, Roni Gillis, Daniela Katz, Albert Grinshpun

**Affiliations:** Helmsley Cancer Center, Shaare Zedek Medical Center, Jerusalem, Israel; Faculty of Medicine, The Hebrew University of Jerusalem, Jerusalem, Israel; The Rachel and Selim Benin School of Computer Science and Engineering, The Hebrew University of Jerusalem, Jerusalem, Israel

**Author notes:** **Corresponding author:** Adar Yaacov, MD.

**Keywords:** CBFB, endocrine therapy, breast cancer, predictive biomarker, MSK-CHORD, METABRIC

## Abstract

**Purpose:** Beyond estrogen receptor (ER) positivity, no genomic biomarker reliably identifies ER+ breast cancer patients who derive differential benefit from endocrine therapy (ET). We performed an unbiased genomic screen to discover genes predicting ET response and characterized the top candidate across clinical settings, treatment modalities, and an independent validation cohort.

**Experimental Design:** We screened 240 genes in 1,197 metastatic ET-treated patients from the MSK-CHORD clinical genomics database using Cox proportional hazards regression with false discovery rate (FDR) correction. The top candidate, core-binding factor subunit beta (CBFB), was characterized across four cohorts defined by disease setting (metastatic/adjuvant) and treatment (ET/chemotherapy), with multivariable adjustment, gene-by-treatment interaction testing, left-truncation sensitivity analysis for guarantee-time bias, and external validation in METABRIC (N = 1,499 ER+).

**Results:** CBFB mutations (prevalence, ∼5%) were the only gene associated with improved time to progression (TTP). In metastatic ET patients, CBFB-mutated tumors (n = 80) demonstrated significantly longer TTP (hazard ratio [HR], 0.44; 95% CI, 0.29–0.67; P = .0002, FDR q = .010) with no chemotherapy benefit (HR, 1.16; P = .65). The gene-by-treatment interaction was significant (HR, 0.37; P = .009). Effects were robust to multivariable adjustment (HR, 0.46–0.50), independent of histology, and preserved under left-truncated Cox regression (HR, 0.38). In the adjuvant setting, CBFB mutations predicted improved recurrence-free survival (HR, 0.52; 95% CI, 0.31–0.85; P = .010), with no effect under chemotherapy. In METABRIC, CBFB mutations predicted improved ER+ overall survival (HR, 0.52; P = 9.3 × 10^−5^).

**Conclusions:** CBFB mutations identify ∼5% of ER+ breast cancers with exceptional ET benefit. As CBFB is included on all major cancer gene panels, this biomarker requires no additional testing infrastructure for clinical implementation.

**CBFB Translational Relevance:** Endocrine therapy is the standard of care for estrogen receptor (ER)–positive breast cancer. Yet beyond ER positivity, clinicians lack genomic biomarkers that identify patients with exceptional response to endocrine treatment. We report that core-binding factor subunit beta (CBFB) mutations predict an extended duration of response to endocrine therapy (hazard ratio, 0.44; P = .0002) with no chemotherapy benefit (hazard ratio, 1.16; P = .65). The gene-by-treatment interaction (P = .009) formally establishes CBFB as a predictive rather than a prognostic biomarker, and the survival association was independently validated in the METABRIC cohort (hazard ratio, 0.52; P = 9.3 × 10^−5^). Because CBFB is already sequenced on major cancer gene panels in clinical use, no additional testing infrastructure is required: CBFB mutation status is a low-cost candidate for prospective validation in endocrine-therapy studies, and, if confirmed, could be reported alongside existing panel results to inform treatment decisions.

## Introduction

Estrogen receptor–positive (ER+) breast cancer accounts for approximately 70% of all breast cancers and is treated primarily with endocrine therapy (ET), including tamoxifen, aromatase inhibitors, and selective endocrine receptor degraders (SERD) like fulvestrant^1,2^. While ET substantially improves outcomes across disease settings, heterogeneity in treatment response remains a central clinical challenge. In the metastatic setting, 30% to 40% of patients demonstrate primary resistance to first-line ET, and the majority of initial responders eventually acquire resistance to the treatment^3,4^. In the adjuvant setting, 20% to 30% of patients experience disease recurrence despite years of ET^4^. Despite these well-recognized limitations, no positive genomic biomarker is used routinely to predict which patients will derive differential benefit from ET. Existing markers predict resistance rather than sensitivity; ESR1 activating mutations represent the most established, predicting resistance to aromatase inhibitors and guiding selection of SERDs^5,6^. Other existing tools such as the Oncotype DX 21-gene recurrence score^7^ provide prognostic and predictive information but do not identify hyper responders to endocrine treatment ^8,9^.

CBFB encodes core-binding factor subunit beta, the non-DNA-binding regulatory partner of the RUNX family of transcription factors (RUNX1, RUNX2, RUNX3). The CBFβ heterodimeric complex plays essential roles in hematopoiesis, osteogenesis, and epithelial differentiation^10,11^. In breast cancer, mutations in CBFB occur in approximately 4% to 6% of tumors^12^. Ciriello et al identified CBFB mutations enriched in invasive lobular carcinoma (ILC)^13^, and subsequent genomic studies have confirmed CBFB as a recurrently altered gene in breast cancer^14,15^. Previous work has established molecular connections between the CBF complex and ER signaling: RUNX1 functions as an ERα tethering factor^16^, controls ER-positive luminal cell fate^17^, and the CBFB–RUNX1 complex suppresses NOTCH signaling in breast cancer^18^. These connections enable a link between CBFB loss-of-function and altered ET response biologically plausible. However, no study to date has investigated whether CBFB mutations predict treatment response or carry any treatment-specific clinical significance.

In this study, we performed an unbiased screen of cancer related genes across the MSK-CHORD dataset^19^ and identified CBFB loss-of-function mutations as the sole gene associated with improved time to progression on ET (FDR q = 0.010). We then characterized this finding across disease settings, treatment modalities, and histological subgroups, assessed robustness through multivariable modeling and sensitivity analyses addressing guarantee-time bias, and validated the survival association externally in the METABRIC cohort^20^.

## Materials and Methods

### Study Populations and Cohort Construction

Patients were identified from the MSK-CHORD dataset, a comprehensive clinical genomics database integrating tumor sequencing, electronic health records, and natural language processing–derived clinical features from Memorial Sloan Kettering Cancer Center^19^. We selected breast cancer patients with available genomic data and first-line treatment information. Patients were classified as metastatic or non-metastatic/adjuvant based on documented metastatic sites.

Within each setting, patients were categorized by first-line treatment: endocrine therapy (ET; hormone therapy excluding CDK4/6 inhibitor combinations) or chemotherapy. To ensure clean treatment comparisons, patients who received both ET and chemotherapy within the same setting were excluded. This yielded four non-overlapping analysis cohorts: metastatic ET, metastatic chemotherapy, adjuvant ET, and adjuvant chemotherapy. Within each setting, the ET and chemotherapy cohorts were mutually exclusive by design. The complete patient flow is presented in Figure S1. Tumors were classified by OncoTree code into invasive ductal carcinoma (IDC), invasive lobular carcinoma (ILC), and other subtypes.

### Genomic Data and CBFB Mutation Definitions

Tumor DNA was sequenced using the MSK-IMPACT platform, a hybridization capture–based next-generation sequencing assay targeting cancer-associated genes^21,22^. All IMPACT-panel genes with ≥5 mutated patients in the metastatic ET cohort were screened. CBFB mutation status was defined as binary (mutated vs. wild-type), including all non-silent mutations detected in the gene. Mutations were classified by functional consequence: loss-of-function (LoF; splice-site, nonsense, frameshift, translation start site), missense, and in-frame deletions. All missense variants were annotated using OncoKB^23^, and mutations were mapped to protein domains to assess distribution across the coding sequence. Mutation-definition sensitivity analyses (loss-of-function only; missense only; hotspot vs non-hotspot) are reported in Supplementary Figure S10 and confirm the primary signal is not driven by a single mutation class.

### Endpoints

Endpoint definition differed by clinical setting to match the biology of each cohort. In the metastatic cohorts, the primary endpoint was time to progression (TTP) — time from first-line treatment initiation to NLP-detected radiographic progression, with censoring at next-line treatment, death, or last follow-up. In the adjuvant cohorts, the primary endpoint was recurrence-free survival (RFS) — time from adjuvant therapy initiation to NLP-detected progression (recurrence) or death from any cause, censoring at last follow-up rather than at planned treatment-line changes (e.g., tamoxifen → AI sequence). This avoids inflating apparent event rates by counting planned regimen switches as administrative censoring. Overall survival (OS) was the secondary endpoint across all cohorts.

### Statistical Analysis

#### Gene-level discovery screen

We performed an unbiased screen of all genes with at least 5 mutations in the metastatic ET cohort (N = 240 IMPACT-panel genes after this prevalence filter) using univariate Cox proportional hazards regression for TTP. P values were corrected using the Benjamini–Hochberg FDR procedure, with significance defined at FDR < 0.05. This screen was pre-specified to identify candidate genes warranting detailed characterization.

#### Multivariable analyses

To assess robustness, multivariable Cox proportional hazards models were fitted in a nested hierarchy: Model A (CBFB + age), Model B (CBFB + age + ILC status), and Model C (CBFB + age + ILC + TP53 + ESR1). Age and histology were selected as covariates because they represent pre-existing patient characteristics that may confound the CBFB–outcome association. Adjusting for mediators biases effect estimates toward the null^24^.

#### Interaction testing

To formally test whether a given biomarker is predictive (treatment-specific) rather than a prognostic biomarker, we combined the ET and chemotherapy cohorts within each setting and fitted the interaction model: h(t) = h_0_(t) × exp(β_1_ × biomarker + β_2_ × IS_ET + β_3_ × biomarker × IS_ET + β_4_ × Age + β_5_ × IS_ILC). The coefficient β_3_ represents the differential effect of the biomarker in ET versus chemotherapy. A significant interaction indicates treatment-specific (predictive) rather than treatment-independent (prognostic) behavior.

#### Left-truncation sensitivity analysis

MSK-CHORD is a clinical sequencing cohort in which patients must survive to the sequencing date to enter the study. For patients whose treatment started before sequencing, person-time between treatment initiation and sequencing is “immortal” – the patient cannot experience an observable event during this interval, a potential source of guarantee-time bias^25,26^. We addressed this by performing left-truncated (delayed-entry) Cox regression, setting each patient’s entry time to the sequencing date rather than the treatment start date. Patients whose treatment ended before sequencing were excluded entirely. As independent confirmation, we also analyzed the post-sequencing-only subset, who have zero immortal time by definition.

#### Power calculations

The Schoenfeld formula for Cox proportional hazards regression was used to assess whether non-significant results in validation subgroups reflected true null effects or insufficient statistical power^27^.

#### Proportional hazards assumption

The proportional hazards assumption was assessed using Schoenfeld residual tests for all primary Cox models.

All P values were two-sided with significance at α = .05. The primary discovery screen used FDR correction. Subsequent characterization analyses within the ET cohorts (interaction testing, multivariable models, histology subgroups) were pre-specified and are presented without additional correction, with effect sizes and confidence intervals reported. Analyses were performed in Python 3.12 using lifelines (survival analysis), scipy (statistical tests), and pandas (data manipulation).

### External Validation in METABRIC

External validation was performed using the Molecular Taxonomy of Breast Cancer International Consortium (METABRIC) dataset^20^, accessed via cBioPortal. The METABRIC cohort comprises 2,509 breast cancer patients with comprehensive genomic profiling and long-term clinical follow-up. We analyzed the ER+ subset for OS and relapse-free survival (RFS), and the ER+ IDC hormone-treated subset for subgroup analysis.

### Structural Analysis of CBFB Variants

Somatic CBFB missense variants identified in MSK-CHORD and germline RUNX1 missense variants from ClinVar (accessed March 2026) were mapped onto solved structures of the CBFB–RUNX1 Runt domain–DNA ternary complex (PDB: 1H9D and 1IO4)^10,28^. ClinVar variants were restricted to those classified as “Pathogenic” or “Likely Pathogenic” with at least a one-star review status^29^. Two functional interfaces were defined: (i) the RUNX1 Runt domain–DNA contact surface and (ii) the RUNX1–CBFB heterodimerization interface. RUNX1–CBFB interface residues were defined using PIONEER^30^, a state-of-the-art protein–protein interface (PPI) prediction framework that integrates evidence from experimentally resolved structures, homology models, and the PIONEER deep-learning model. Residues predicted to participate in a PPI by any of these sources were considered interface positive. Enrichment of somatic CBFB missense variants at interface-positive versus non-interface positions was assessed using Fisher’s exact test.

## Data Availability

This study used de-identified, publicly available data from the MSK-CHORD cohort, accessed through cBioPortal (https://www.cbioportal.org/study/summary?id=msk_chord_2024). The primary MSK-CHORD study was approved by the Memorial Sloan Kettering Cancer Center Institutional Review Board for retrospective analysis of de-identified clinical data

## Data Availability

The MSK-CHORD dataset is available through cBioPortal (https://www.cbioportal.org/study/summary?id=msk_chord_2024). The METABRIC dataset is available through cBioPortal (https://www.cbioportal.org/study/summary?id=brca_metabric).

## Code Availability

Analysis code is available at https://github.com/adarya/cbfb-predictive.

## Ethics Statement

This study used de-identified, publicly available data from the MSK-CHORD cohort, accessed through cBioPortal (https://www.cbioportal.org/study/summary?id=msk_chord_2024). The primary MSK-CHORD study was approved by the Memorial Sloan Kettering Cancer Center Institutional Review Board for retrospective analysis of de-identified clinical data. Secondary analysis of this publicly available, de-identified dataset did not require additional institutional review board approval at the authors’ institutions, in accordance with local regulations governing research on de-identified public data.

## Results

### Cohort Characteristics

The study included 5,191 breast cancer patients from the MSK-CHORD dataset (Table 1). Patients were classified as metastatic (N = 3,793) or adjuvant (N = 1,939); 541 appeared in both settings due to disease progression. After excluding patients who received both ET and chemotherapy within the same setting (metastatic: n = 1,209; adjuvant: n = 274), the four analysis cohorts comprised: metastatic ET (N = 1,197), metastatic chemotherapy (N = 995), adjuvant ET (N = 861), and adjuvant chemotherapy (N = 566). The histological distribution was IDC (3,233; 62.3%), ILC (525; 10.1%), and other subtypes (1,433; 27.6%). CBFB mutations were present in 240 patients overall (4.6%), with significant enrichment in ILC (9.1% vs. 4.1% in IDC; odds ratio, 2.34; P = 3.6 × 10^−7^; Table ST1).

**Table 1.** Cohort Characteristics.

| Characteristic | Overall (N=5,191) | Met ET (N=1,197) | Met Chemo (N=995) | Adj ET (N=861) | Adj Chemo (N=566) |
| --- | --- | --- | --- | --- | --- |
| IDC | 3,233 (62.3%) | 731 (61.1%) | – | 555 (64.5%) | – |
| ILC | 525 (10.1%) | 159 (13.3%) | – | 95 (11.0%) | – |
| Other | 1,433 (27.6%) | 307 (25.6%) | – | 211 (24.5%) | – |
| CBFB Mutated | 240 (4.6%) | 80 (6.7%) | 19 (1.9%) | 54 (6.3%) | 29 (5.1%) |
| CBFB Wild-type | 4,951 (95.4%) | 1,117 (93.3%) | 976 (98.1%) | 807 (93.7%) | 537 (94.9%) |
| Age, median (yr) | – | 60 | – | 65 | – |

Abbreviations: Met, metastatic; Adj, adjuvant; ET, endocrine therapy; IDC, invasive ductal carcinoma; ILC, invasive lobular carcinoma.

### An Unbiased Genomic Screen Identifies Association of CBFB with Improved Endocrine Therapy Outcomes

We screened 240 genes in 1,197 metastatic ET-treated patients (all IMPACT-panel genes with ≥5 mutated patients in the metastatic ET cohort; see Methods). Eight genes reached FDR significance for association with TTP (Figure 1A,B). Seven predicted shorter TTP and included well-established resistance-associated genes: TP53 (HR, 1.74; FDR q < .001), ESR1 (HR, 1.96; q < .001), and RB1 (HR, 2.09; q = .012). CBFB was the only FDR-significant gene associated with improved TTP (HR, 0.44; FDR q = .010; rank 3 of 240), making it the sole positive predictive candidate to emerge from the screen. In metastatic ET-treated patients, CBFB-mutated tumors (n = 80) demonstrated significantly longer TTP compared with wild-type tumors (HR, 0.44; 95% CI, 0.29–0.67; P = .0002; Figure 1C). In contrast, CBFB mutations conferred no benefit in the metastatic chemotherapy cohort (HR, 1.16; 95% CI, 0.62–2.16; P = .65; Figure 1D). Overall survival showed a concordant pattern: metastatic ET HR, 0.49 (P = .012; Figure S2A); metastatic chemotherapy HR, 1.40 (P = .25; Figure S2B).

**Figure 1.**
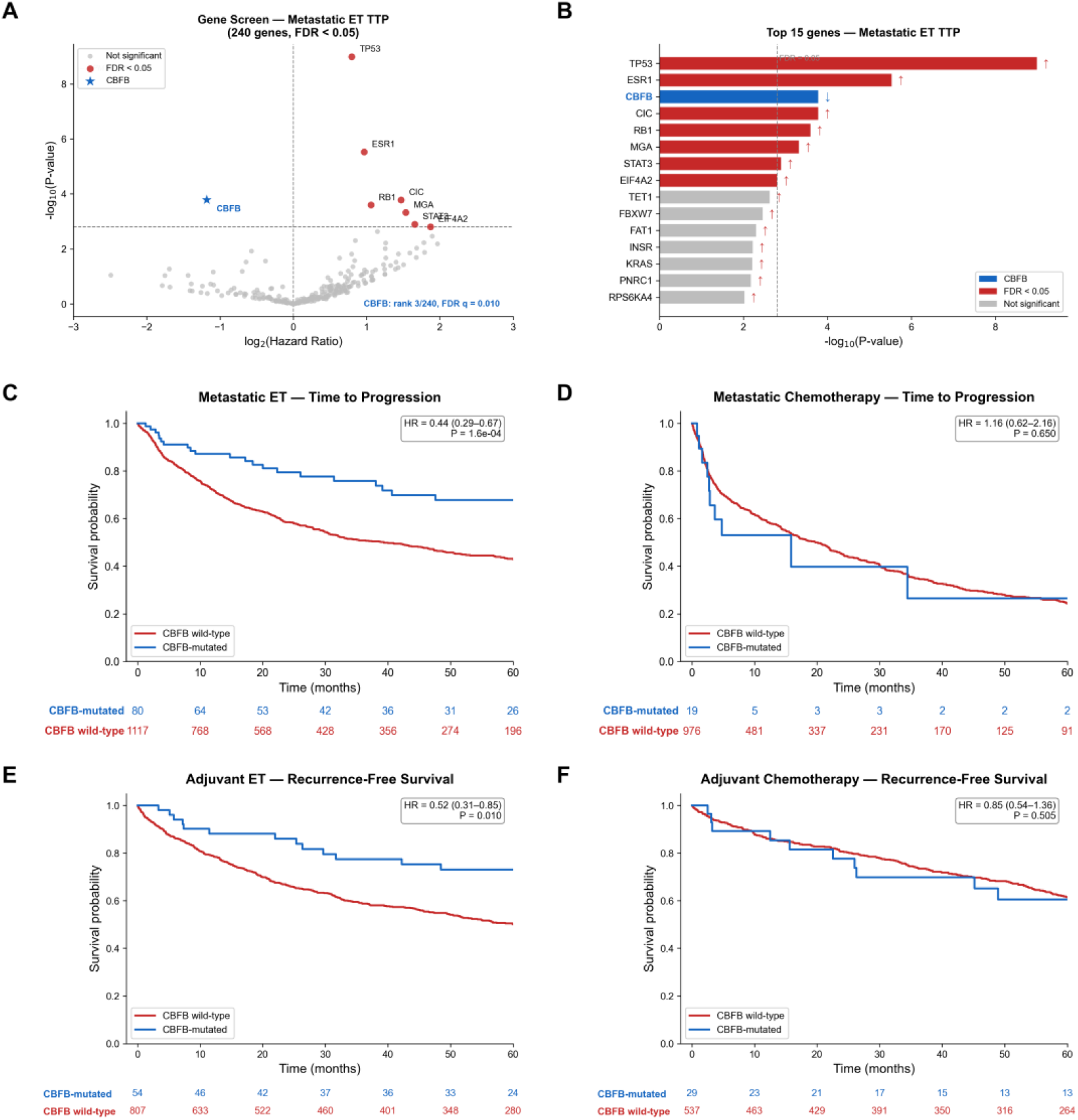
Unbiased genomic screen identifies CBFB as the sole gene predicting endocrine therapy benefit. (A-B) Volcano plot of the 240-gene discovery screen in metastatic ET TTP and barplot of the top 15 genes. (C–D) Kaplan–Meier curves for time to progression in the metastatic ET and metastatic chemotherapy cohorts. (E–F) Kaplan–Meier curves for recurrence-free survival in the adjuvant ET and adjuvant chemotherapy cohorts (RFS used in lieu of TTP for adjuvant settings to avoid administrative censoring at planned regimen changes): (C) Metastatic ET (N = 1,197). (D) Metastatic chemotherapy (N = 995). (E) Adjuvant ET (N = 861). (F) Adjuvant chemotherapy (N = 566). P values from univariate Cox proportional hazards regression. Numbers at risk are shown below each Kaplan–Meier curve.

This treatment-specific pattern was replicated in the adjuvant setting. CBFB mutations predicted benefit from adjuvant ET (RFS: HR, 0.52; 95% CI, 0.31–0.85; P = .010; Figure 1E) with no effect in the adjuvant chemotherapy cohort (RFS HR, 0.85; 95% CI, 0.54–1.36; P = .505; Figure 1F). Adjuvant OS results were consistent in direction (ET: HR, 0.50; P = .032; chemotherapy: HR, 1.10; P = .73, Figure S2C,D). Across all four cohorts, the pattern was unambiguous: significant benefit restricted to ET, with null effects under chemotherapy (Table 2).

**Table 2.** CBFB Mutation Association with Outcomes across Cohorts.

| Setting | Endpoint | Treatment | Subgroup | N | n_mut | HR | 95% CI | P |
| --- | --- | --- | --- | --- | --- | --- | --- | --- |
| Metastatic | TTP | ET | All | 1,197 | 80 | 0.44 | 0.29–0.67 | .0002 |
| Metastatic | TTP | Chemo | All | 995 | 19 | 1.16 | 0.62–2.16 | .65 |
| Adjuvant | RFS | ET | All | 861 | 54 | 0.52 | 0.31–0.85 | .010 |
| Adjuvant | RFS | Chemo | All | 566 | 29 | 0.85 | 0.54–1.36 | .505 |
| Metastatic | OS | ET | All | 1,197 | 80 | 0.49 | 0.28–0.85 | .012 |
| Metastatic | OS | Chemo | All | 995 | 19 | 1.40 | 0.79–2.49 | .25 |
| Adjuvant | OS | ET | All | 861 | 54 | 0.50 | 0.27–0.94 | .032 |
| Adjuvant | OS | Chemo | All | 566 | 29 | 1.10 | 0.63–1.92 | .73 |
| Metastatic | TTP | ET | IDC | 731 | 46 | 0.38 | 0.20–0.71 | .003 |
| Metastatic | TTP | ET | ILC | 159 | 20 | 0.30 | 0.09–0.95 | .041 |
| Metastatic | OS | ET | IDC | 731 | 46 | 0.31 | 0.13–0.76 | .011 |
| Adjuvant | RFS | ET | IDC | 555 | 41 | 0.52 | 0.29–0.96 | .037 |
| Adjuvant | OS | ET | IDC | 555 | 41 | 0.39 | 0.16–0.95 | .039 |
| Metastatic | TTP | – | Adj. | 2,192 | 99 | 0.37 | 0.17–0.78 | .009 |
| Metastatic | OS | – | Adj. | 2,192 | 99 | 0.36 | 0.16–0.79 | .011 |
| Adjuvant | RFS | – | Adj. | 1,427 | 83 | 0.60 | 0.31–1.19 | .145 |
| Adjuvant | OS | – | Adj. | 1,427 | 83 | 0.46 | 0.20–1.07 | .070 |

NOTE: Bold indicates P < .05. Interaction models adjusted for age and ILC status. Abbreviations: HR, hazard ratio; CI, confidence interval; TTP, time to progression; RFS, recurrence free survival; OS, overall survival; ET, endocrine therapy; Chemo, chemotherapy; Adj., adjusted for age and ILC; IDC, invasive ductal carcinoma; ILC, invasive lobular carcinoma.

The CBFB–ET association was robust to sequential covariate adjustment (Figure 2A; Table ST2). Age-adjusted model yielded HR = 0.44 (P = .0002). Adding ILC status modestly yielded HR = 0.46 (95% CI, 0.30–0.70; P = .0004). The most stringent model, further adjusting for TP53 and ESR1, yielded HR = 0.50 (95% CI, 0.33–0.77; P = .002).

**Figure 2.**
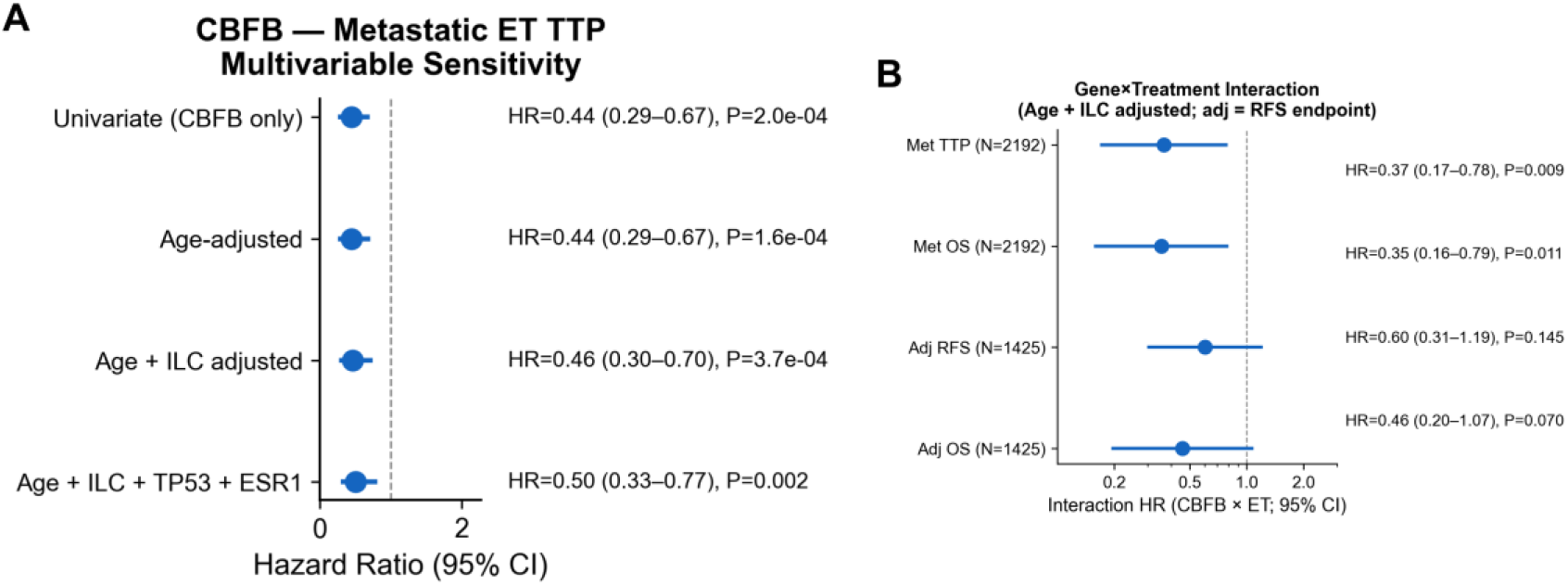
CBFB association is robust to multivariable adjustment and treatment-specific by formal interaction testing. (A) Forest plot of CBFB hazard ratios across nested multivariable models in the metastatic ET cohort (univariate, age-adjusted, age + ILC-adjusted, and age + ILC + TP53 + ESR1-adjusted). (B) Forest plot of age- and ILC-adjusted gene-by-treatment interaction hazard ratios across disease settings and endpoints (Met TTP, Met OS, Adj RFS, Adj OS). Interaction HR < 1 indicates a treatment-specific (predictive) biomarker for endocrine therapy.

### Gene-by-Treatment Interaction Establishes CBFB as a Predictive Biomarker

In the metastatic setting, the CBFB × ET interaction was significant for both TTP (interaction HR, 0.37; 95% CI, 0.17–0.78; P = .009) and OS (interaction HR, 0.36; 95% CI, 0.16–0.79; P = .011), formally establishing that CBFB mutations predict differential treatment benefit rather than a treatment-independent favorable prognosis (Figure 2B). In the adjuvant setting, the interaction was directionally consistent under both endpoints but did not reach formal significance (RFS: HR, 0.60; 95% CI, 0.31–1.19; P = .145; OS: HR, 0.46; P = .070), as expected given the smaller adjuvant sample size and lower event rate.

### CBFB Effect Is Independent of Histological Subtype

Because CBFB mutations are enriched in ILC (OR, 2.34), a critical question is whether the observed ET benefit is driven by ILC-associated favorable biology. Three lines of evidence demonstrate histological independence. First, the majority of CBFB-mutated ET patients had IDC histology: 46 of 80 (57.5%) in the metastatic ET cohort and 87 of 134 (64.9%) combined across ET cohorts (Figure 3A). Second, CBFB predicted ET benefit within IDC alone: among metastatic ET-treated IDC patients (N = 731), those with CBFB mutations (n = 46) demonstrated significantly longer TTP (HR, 0.38; 95% CI, 0.20–0.71; P = .003; Figure 3B) and OS (HR, 0.31; P = .011). Third, multivariable regression adjusting for age and ILC confirmed CBFB as an independent predictor (HR, 0.46; P = .0004).

**Figure 3.**
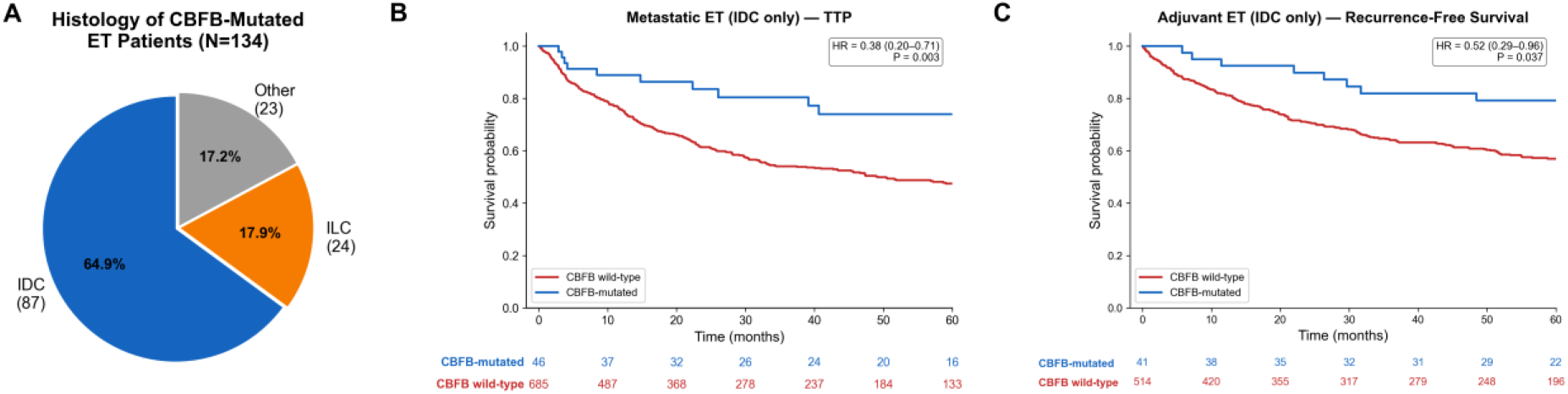
CBFB effect is independent of histological subtype. (A) Pie chart of histological distribution among CBFB-mutated ET patients across both settings (N = 134. (B,C) Kaplan–Meier curves for within IDC-only patients: (B) TTP in metastatic ET IDC subset (N = 731); (C) RFS in adjuvant ET IDC subset (N = 555. Numbers at risk are shown below each Kaplan–Meier curve.

### CBF Complex–Level Analysis

RUNX1 alone showed a concordant but borderline association in the metastatic ET screen (rank 40 of 240; HR, 0.62; P = .065; n = 45). Combining CBFB and RUNX1 mutations as a single CBF-complex biomarker (n = 122 in metastatic ET) yielded a highly significant TTP association (HR, 0.48; 95% CI, 0.34–0.67; P = 2.0 × 10^−5^; Table ST3). Treatment specificity was preserved: metastatic chemotherapy TTP HR = 0.94 (P = .76). In the adjuvant setting, the CBF-complex biomarker similarly predicted ET benefit (RFS HR, 0.60; 95% CI, 0.41–0.88; P = .009; Figure 4A,B).

**Figure 4.**
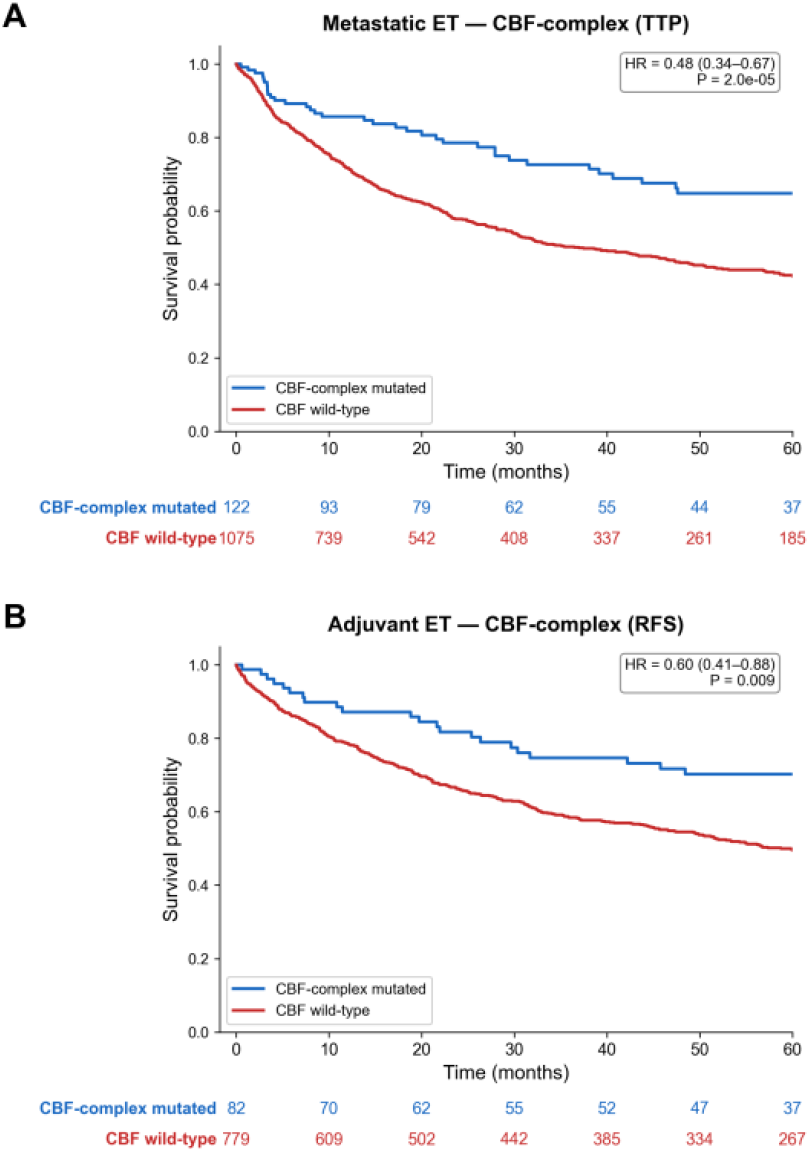
CBF-complex (CBFB or RUNX1) mutations recapitulate the treatment-specific endocrine sensitivity signal. (A) Kaplan–Meier curve for TTP in the metastatic ET cohort, stratified by CBF-complex (CBFB or RUNX1) mutation status (N = 1,197). (B) Kaplan–Meier curve for RFS in the adjuvant ET cohort, stratified by CBF-complex mutation status (N = 861). Numbers at risk are shown below each Kaplan–Meier curve. Chemotherapy negative controls remain null (Met chemo TTP CBF-complex HR, 0.94; P = .76).

As an exploratory co-mutation analysis, the CBFB × PIK3CA interaction was significant for TTP in metastatic ET (interaction HR, 0.21; 95% CI, 0.08–0.58; P = .003; Figure S3), with the joint CBFB/PIK3CA-mutated subgroup showing the strongest endocrine-sensitivity signal (HR, 0.17; 95% CI, 0.07–0.42; P < .001; Figure S4). This cooperative pattern is hypothesis-generating and warrants prospective validation. The CBFB-mutated subset showed significant co-occurrence with GATA3 (FDR q = 1 × 10^−5^), CDH1 (q = .010), and KMT2C (q = .035), and significant mutual exclusivity with TP53 (q = .010; Supplementary Figure S5), consistent with a luminal, ER-axis-intact genomic context.

### External Validation in METABRIC

Among ER+ patients in the METABRIC cohort, CBFB mutations were associated with significantly improved OS (N = 1,499; CBFB-mutated n = 91; HR, 0.52; 95% CI, 0.38–0.72; P = 9.3 × 10^−5^; Figure 5A) and RFS (N = 1,741; n = 103; HR, 0.59; P = .005; Figure 5B). The association was robust within ER+ IDC (OS: HR, 0.49; P = .0006) and was confirmed in the ER+ IDC hormone-treated subset (N = 835; OS: HR, 0.59; 95% CI, 0.36–0.95; P = .031; Figure 5C,D).

**Figure 5.**
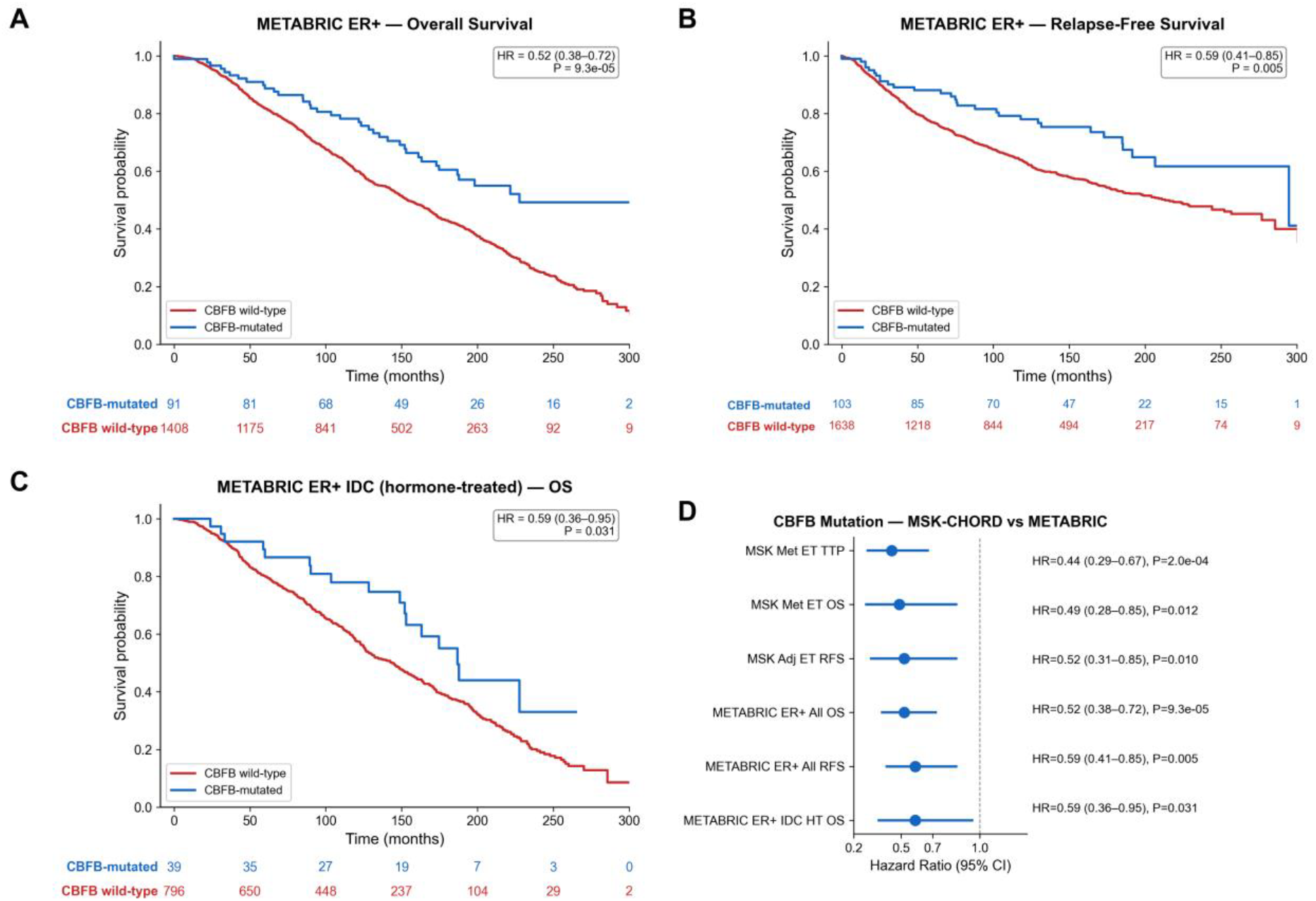
External validation in the METABRIC cohort. (A-B) Kaplan–Meier curve for OS and RFS in METABRIC ER+ patients. (C) Kaplan–Meier curve for overall survival in the METABRIC ER+ IDC hormone-treated subset (N = 835). (D) Forest plot comparing CBFB hazard ratios across MSK-CHORD and METABRIC cohorts. Numbers at risk are shown below the Kaplan–Meier curves in panels A and B.

### Sensitivity Analyses

In the metastatic ET cohort, 59% of patients initiated ET before sequencing, with a median lead time of 150 days. Under left-truncated Cox regression, TTP/RFS hazard ratios were strengthened for both ET cohorts: metastatic ET from 0.44 to 0.38 (P = .001) and adjuvant ET RFS from 0.52 to 0.46 (P = .013; Figure S6). Chemotherapy negative controls remained null under left truncation. The gene-by-treatment interaction was preserved (HR, 0.31; P = .015). In the post-sequencing-only subset (N = 491; CBFB-mutated n = 39), which has zero immortal time by definition, TTP HR was 0.42 (P = .036), replicating the primary finding in a completely unbiased subgroup.

To address whether the CBFB benefit was driven by specific metastatic patterns, we performed subgroup analyses in the metastatic ET cohort by liver-metastasis status and bone-only disease. The CBFB effect was consistent across both dichotomies with no evidence of effect modification (P-interaction = 0.94 for liver, 0.40 for bone-only; Figure S7).

Of 309 CBFB variants across the cohort, 69% were loss-of-function (splice-site n = 87, nonsense n = 47, frameshift insertion n = 36, frameshift deletion n = 35, translation start site n = 7), 29% were missense (n = 90), and 2% were in-frame deletions (n = 7; Figure S8). All missense variants were annotated as “Unknown” by OncoKB, with none classified as oncogenic or likely oncogenic. Ninety-one percent of mutations mapped to the Runt-binding domain (amino acids 1–141), with a single hotspot at splice-site position 55 accounting for 18% of all mutations (Figure S9). The ET benefit was consistent across all tested mutation definitions (Figure S10), with a narrow HR range (0.41–0.44).

### APOBEC mutational signatures

Detecting APOBEC mutational signatures using MESiCA^31^ suggested a potential depletion of APOBEC, a known resistance related mutagenesis mechanism in breast cancer^32^, in the CBFB-mutated tumors. Within the metastatic ET cohort, APOBEC prevalence was directionally lower in CBFB-mutated tumors but did not reach significance (11.2% vs. 14.1%; P = .62). Pooling with the adjuvant ET cohort to increase power preserved the direction (HR+ subset: 11.5% vs. 17.5%; P = .10), a hypothesis-generating trend that warrants confirmation in larger cohorts.

### Structural mapping

RUNX1 and CBFB form an extensive, non-linear interface within the CBF complex (Figure S11A). CBFB does not directly bind DNA but stabilizes the DNA-bound RUNX Runt domain^10,28^, supporting the functional importance of the RUNX1–CBFβ interface. RUNX1 directly contacts DNA through the Runt domain, and pathogenic germline RUNX1 missense variants from ClinVar clustered at the RUNX1–DNA and to a smaller extent at the RUNX1–CBFβ interfaces (Figure S11B). These variants cause RUNX1 familial platelet disorder^33^, an autosomal-dominant thrombocytopenia syndrome with predisposition to hematologic malignancies. The dominant mechanism supports the clinical relevance of single damaging variants affecting CBF-RUNX complex.

Although somatic CBFB missense variants in the MSK-CHORD did not show a single classical hotspot, they were enriched at the RUNX1–CBFB interface (Figure S11C; Fisher exact P = .0030). Five of 10 unique interface variants recurred in more than one patient (R3C, V4G, G61R, Q67H, and M101V), compared with 1 of 18 non-interface variants (P = .126). Somatic RUNX1 variants showed a complementary pattern, with aggregation near DNA-interface residues and overlap with several germline pathogenic positions (Figure S11D). Together, these findings provide structural evidence that somatic CBFB mutations in breast cancer disrupt CBF complex formation and suggest that dedicated structure-aware predictors may improve interpretation of individual CBFB and RUNX1 variants.

## Discussion

This study identifies CBFB mutations as a candidate treatment-specific predictive biomarker for ET benefit in ER+ breast cancer. The finding emerged from an unbiased screen of 240 genes, where CBFB was the only gene associated with improved ET outcomes. The association was consistent across disease settings, treatment-specific by formal interaction testing (P = .009), independent of histology and key genomic covariates, and externally supported by the METABRIC cohort.

The prior literature on CBFB in breast cancer has focused on its role in the molecular landscape of ILC. Ciriello and colleagues reported enrichment of CBFB mutations in lobular carcinoma^13^, and subsequent genomic profiling studies confirmed CBFB as recurrently mutated in breast cancer without investigating treatment response^14,15^. Our study extends this work in two ways. First, the gene-by-treatment interaction formally establishes that the CBFB effect is treatment-specific, not merely a reflection of generally favorable biology. Second, the treatment-predictive effect is independent of ILC histology.

The biological basis for CBFB-mediated ET sensitivity might involve two complementary mechanisms. First, the CBFB–RUNX1 complex directly modulates the ER transcriptional program: RUNX1 serves as an ERα tethering factor that recruits estrogen receptor to specific chromatin loci and controls luminal cell fate through FOXA1 and ELF5. CBFB loss-of-function disrupts RUNX1 stability and transcriptional activity, potentially altering ER-dependent gene expression in ways that sustain ET sensitivity. The structural mapping of somatic CBFB missense variants to the RUNX1-binding interface, the same surface disrupted by germline RUNX1 variants in RUNX1-FPD^33^, provides convergent evidence across somatic and germline contexts for the functional importance of CBF complex integrity. The CBF-complex biomarker analysis supports this pathway-level interpretation, with combined CBFB/RUNX1 mutations retained the predictive effect.

The APOBEC mutagenesis axis offers a candidate resistance-limiting mechanism. APOBEC3B is the dominant source of cytidine deamination–mediated mutagenesis in breast cancer and has been implicated in tamoxifen resistance in ER+ disease^34–36^. A directionally consistent but underpowered trend toward reduced APOBEC enrichment in CBFB-mutated tumors was observed.

Several limitations warrant discussion. First, this is a retrospective analysis of a single-institution clinical sequencing cohort with non-randomized treatment assignment. Unmeasured confounders may influence the results, although the treatment-specificity and consistency across multivariable models argue against simple confounding by indication. Second, guarantee-time bias was addressed through left-truncated Cox regression and analysis of the post-sequencing-only subset; hazard ratios were preserved or strengthened, confirming that naive estimates are conservative. Third, CBFB mutation counts are modest, typical for a gene mutated at approximately 5% frequency and representing the inherent trade-off between specificity and sample size in biomarker discovery. Fourth, whether CBFB predicts benefit from CDK4/6 inhibitor–based regimens remains unknown and is a priority for future investigation.

From the perspective of clinical implementation, CBFB mutations possess an immediately actionable characteristic: the gene is already included on every major cancer gene panel in clinical use, including MSK-IMPACT, FoundationOne CDx, and Guardant360. No additional testing infrastructure, assay development, or regulatory approval is needed to report CBFB mutation status. For the approximately 5% of ER+ breast cancer patients harboring CBFB loss-of-function mutations, this information could be incorporated into existing clinical reports to support treatment decision-making.

In conclusion, CBFB loss-of-function mutations identify approximately 5% of ER+ breast cancers with exceptional ET benefit. The treatment-specific, histology-independent, multivariable-robust association, externally supported by the METABRIC survival data, identifies CBFB as a candidate predictive biomarker warranting prospective investigation. Future directions include prospective validation in ET trials, functional characterization of the CBFB–APOBEC3B mechanistic axis, assessment of CBFB predictive value in the CDK4/6 inhibitor era, and evaluation of circulating tumor DNA–based CBFB detection for real-time treatment monitoring.

## Acknowledgments

We thank the MSK-CHORD team for making the dataset available through cBioPortal. Molecular graphics and analyses were performed with UCSF ChimeraX, developed by the Resource for Biocomputing, Visualization, and Informatics at the University of California, San Francisco, with support from National Institutes of Health R01-GM129325 and the Office of Cyber Infrastructure and Computational Biology, National Institute of Allergy and Infectious Diseases.

## Author Contributions

A.Y.: Conceptualization; Methodology; Software; Formal analysis; Data curation; Visualization; Writing – original draft; Writing – review & editing. G.P.: Investigation; Writing – original draft; Writing – review & editing. R.G.: Methodology; Validation; Writing – review & editing. D.K.: Investigation; Writing – review & editing. A.G.: Conceptualization; Writing – review & editing; Supervision. All authors approved the final manuscript.

## Declaration of generative AI and AI-assisted technologies in the manuscript preparation process

During the preparation of this work the authors used Claude Opus 4.0 and 4.7 in order to assist with analyses, manuscript writing and editing. After using these tools, the authors reviewed and edited the content as needed and take full responsibility for the content of the published article.

