## Supplementary information for "CBFB mutations predict endocrine therapy benefit in estrogen receptor–positive breast cancer"

Contains 11 supplementary figures and 3 supplementary tables.

### Supplementary Figures

**Figure S1. Patient flow diagram (CONSORT-style).**

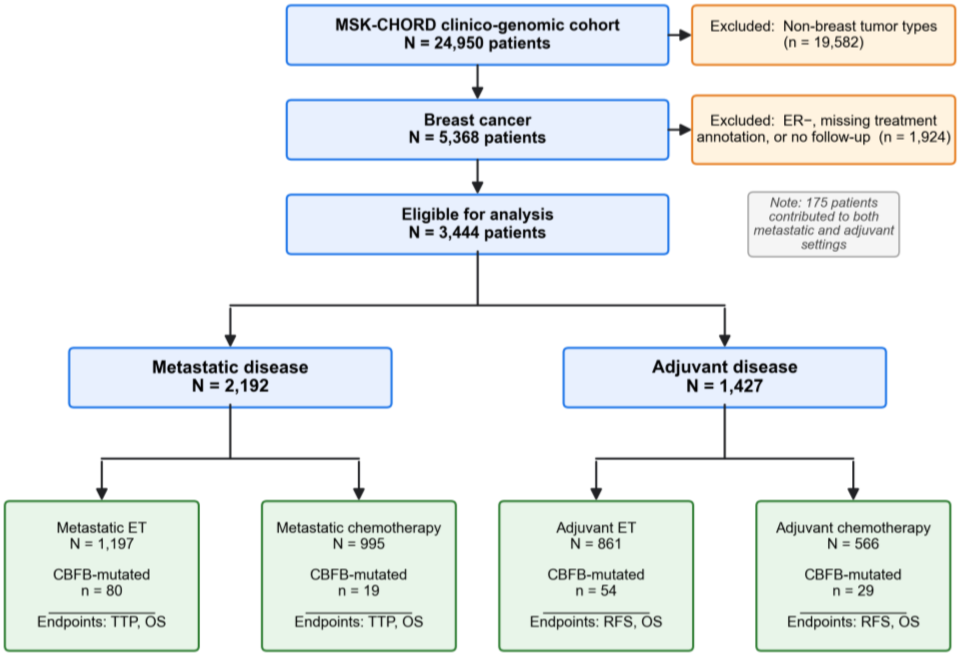


**Figure S1. Patient flow diagram (CONSORT-style).** Patient flow from the MSK-CHORD source population through cohort definition.

**Figure S2. Overall survival by CBFB mutation status in the chemotherapy cohorts (negative controls).**

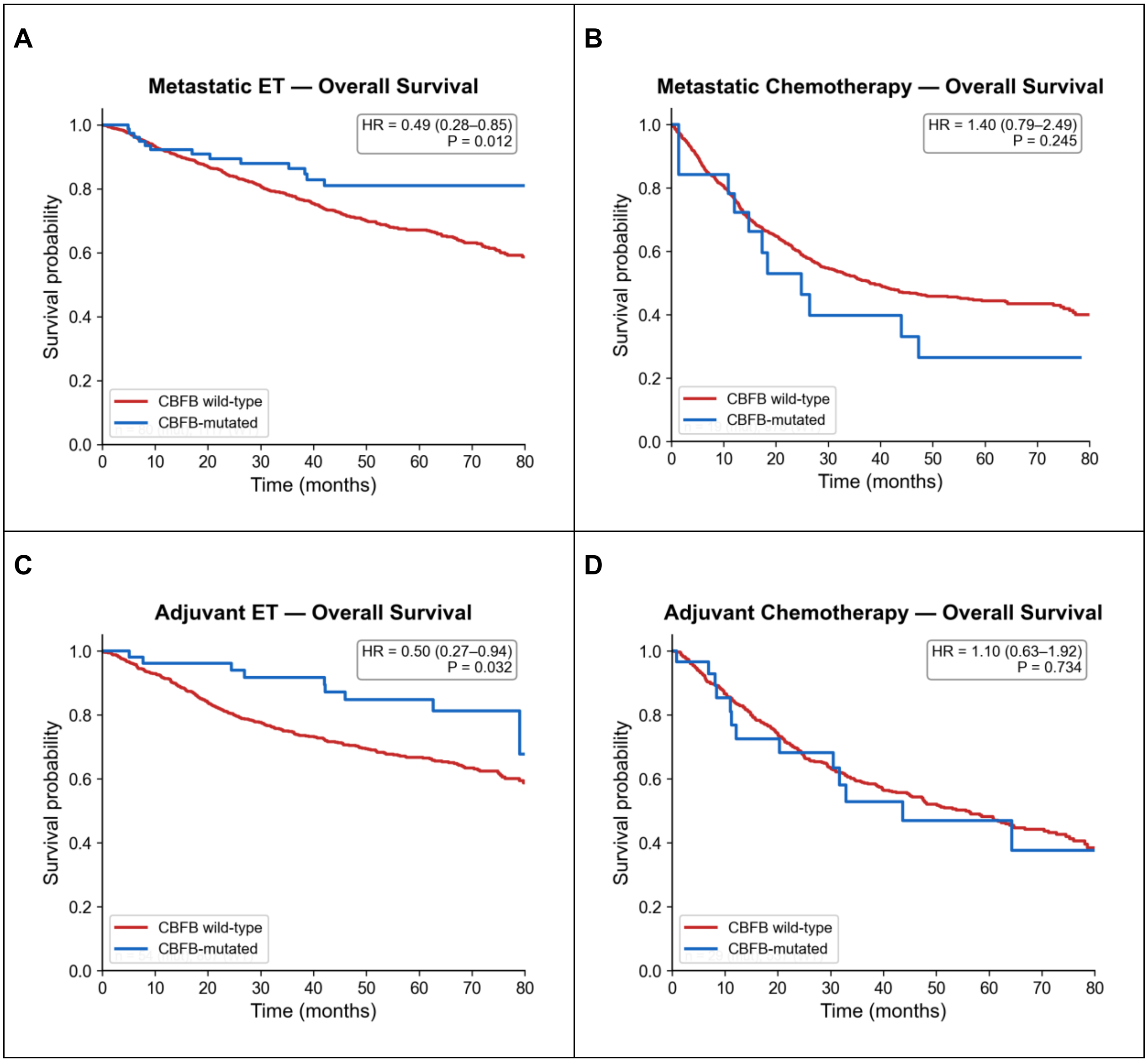


**Figure S2. Overall survival by CBFB mutation status in the chemotherapy cohorts (negative controls).** Kaplan–Meier curves for overall survival (OS) stratified by CBFB mutation status.

**Figure S3. Formal CBFB × PIK3CA interaction test.**

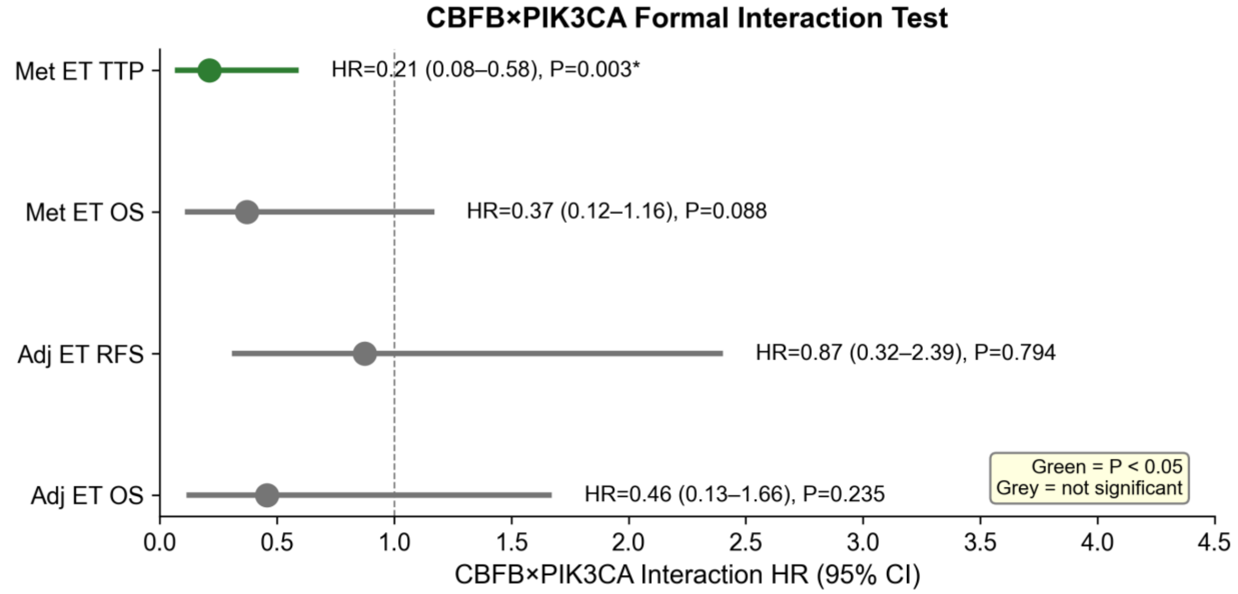


**Figure S3. Formal CBFB × PIK3CA interaction test.** Forest plot of the formal CBFB × PIK3CA interaction model, adjusted for age and ILC status. The significant interaction (P = .003) supports a cooperative effect of CBFB and PIK3CA loss-of-function on ET sensitivity.

**Figure S4. CBF-complex (CBFB + RUNX1) and PIK3CA stratification.**

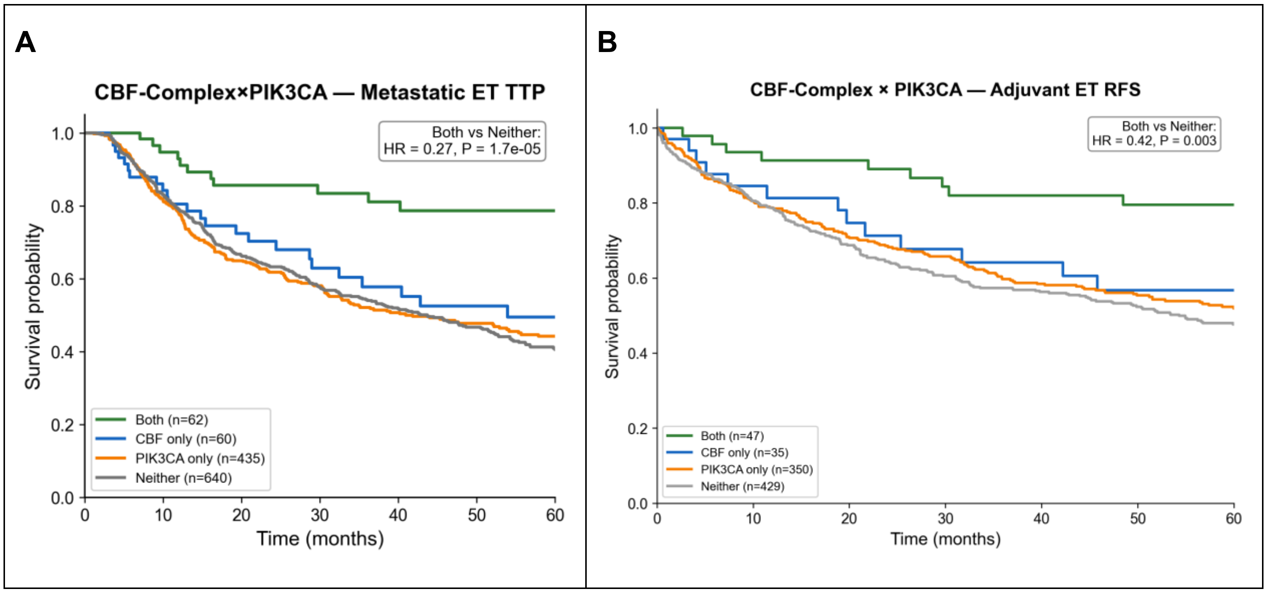


**Figure S4. CBF-complex (CBFB + RUNX1) and PIK3CA stratification.** Kaplan–Meier curves for the joint CBF-complex × PIK3CA stratification, illustrating that the CBF + PIK3CA co-mutation effect generalizes across the CBF complex. (A) Metastatic ET TTP. (B) Adjuvant ET RFS.

#### Figure S5. CBFB co-mutation landscape in the metastatic ET cohort.

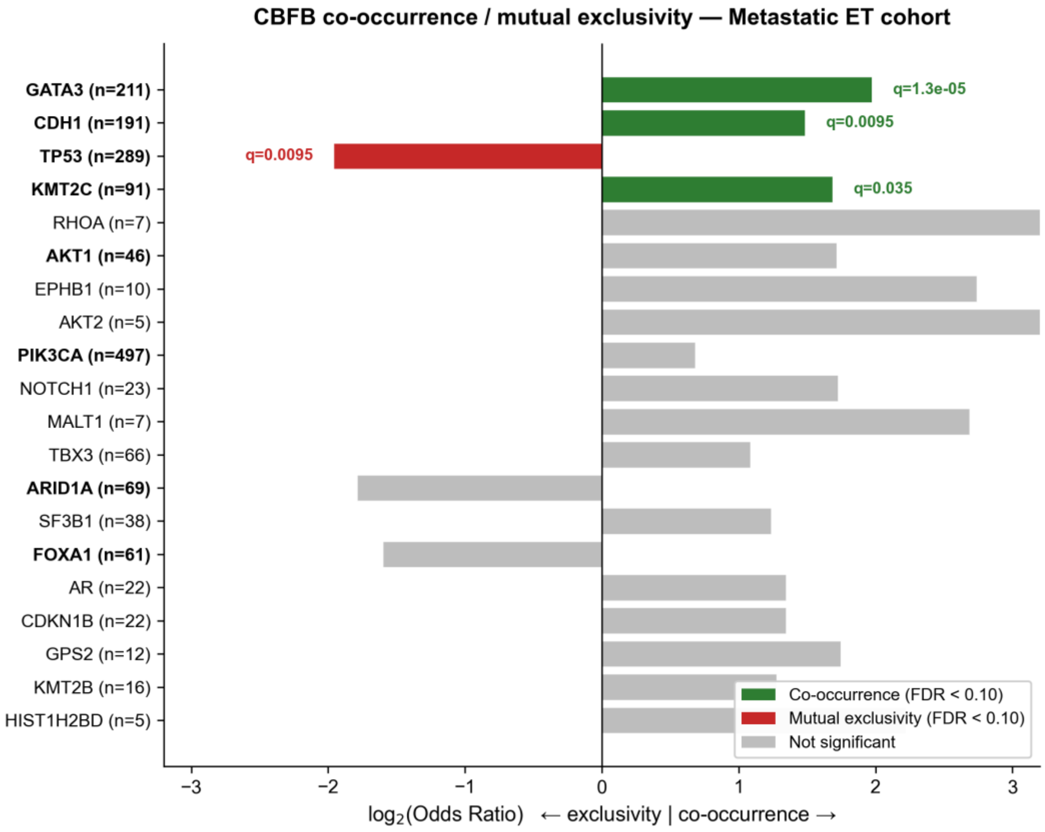


**Figure S5.** CBFB co-mutation landscape in the metastatic ET cohort. Forest of log2 odds ratios (CBFB-mutated vs CBFB–wild-type) for co-occurrence (positive log2 OR) and mutual exclusivity (negative) across all 240 IMPACT-panel genes meeting the prevalence threshold in the metastatic ET cohort. P values are from two-sided Fisher exact tests; FDR correction by Benjamini–Hochberg. Genes reaching FDR q < .05 are labeled (GATA3, CDH1, KMT2C — co-occurring with CBFB; TP53 — mutually exclusive), consistent with a luminal, ER-axis–intact genomic context for the CBFB-mutated subset.

#### Figure S6. Left-truncated Cox sensitivity analysis.

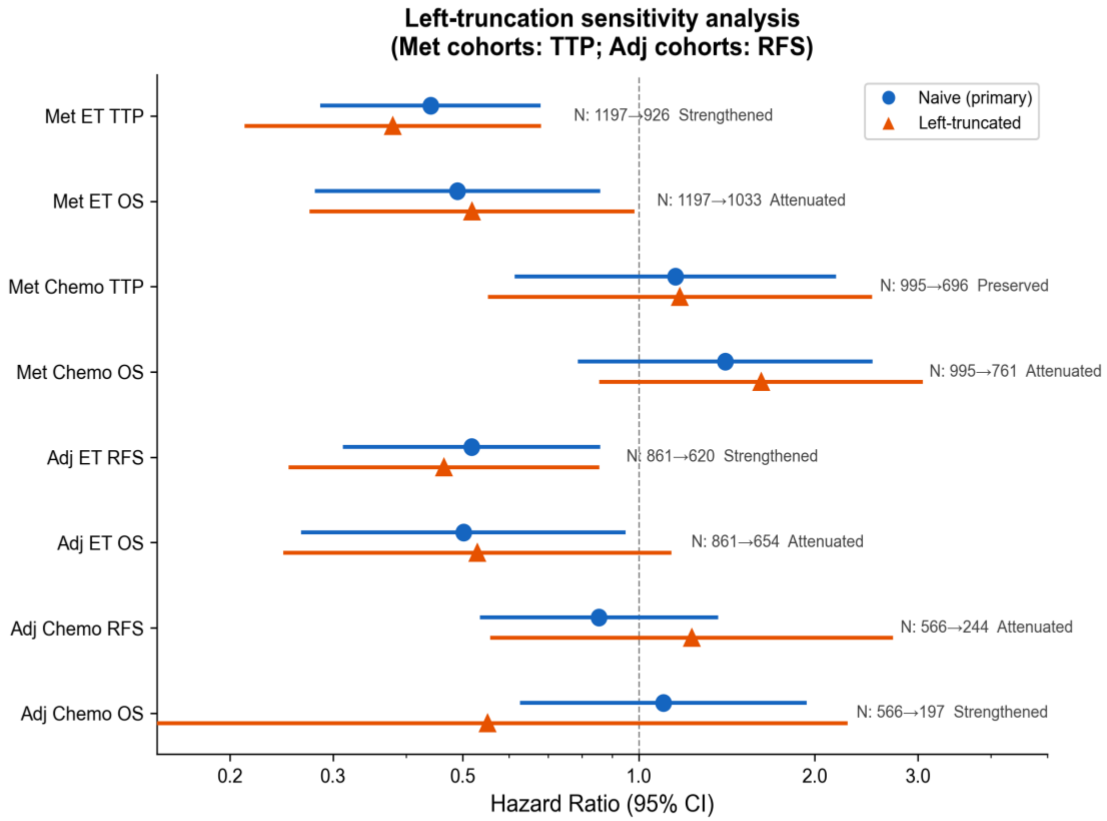


**Figure S6. Left-truncated Cox sensitivity analysis.** Forest plot comparing naive (left-aligned to first treatment) and left-truncated (delayed-entry at sequencing date) Cox regression hazard ratios across the four cohorts. Hazard ratios for the ET cohorts are preserved or strengthened under left truncation, indicating that the primary findings are not driven by guarantee-time bias.

**Figure S7. Subgroup analysis: CBFB effect on TTP in metastatic ET by metastatic pattern.**

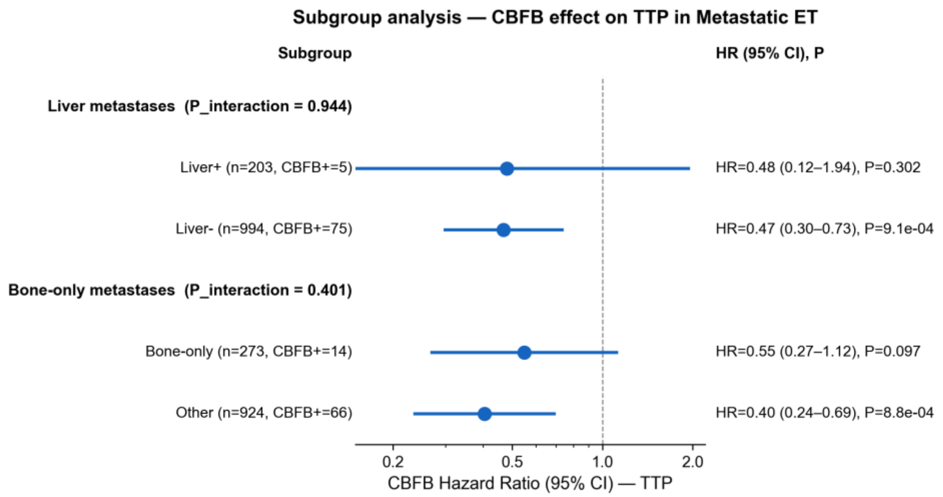


**Figure S7. Subgroup analysis: CBFB effect on TTP in metastatic ET by metastatic pattern.** Forest plot of CBFB hazard ratios for time to progression in the metastatic ET cohort stratified by (top) liver metastasis status and (bottom) bone-only disease. Subgroup-level HRs with 95% CI and P values are shown; CBFB × subgroup interaction P values test for effect modification. No significant heterogeneity was observed (liver P_interaction = 0.94; bone-only P_interaction = 0.40), supporting that the CBFB benefit generalizes across the dominant metastatic-presentation patterns.

**Figure S8. CBFB mutation spectrum.**

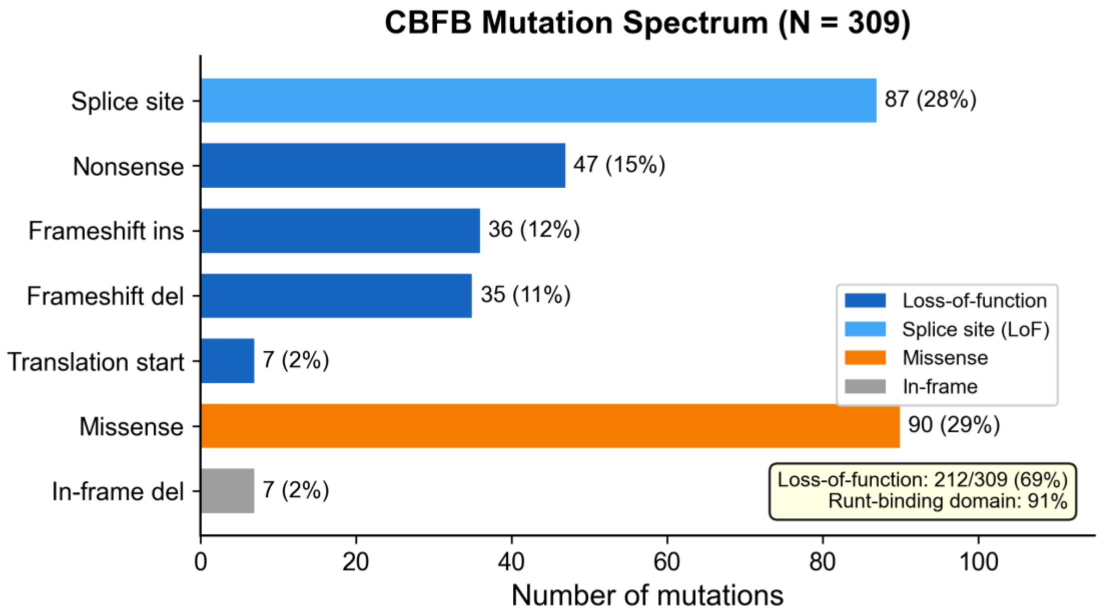


**Figure S8. CBFB mutation spectrum.** Distribution of CBFB variant classes across the cohort (N = 309 variants in 240 patients). Loss-of-function variants (splice-site, nonsense, frameshift insertion/deletion, translation start site) account for 69% of variants; missense for 29%; in-frame deletions for 2%.

**Figure S9. CBFB lollipop plot.**

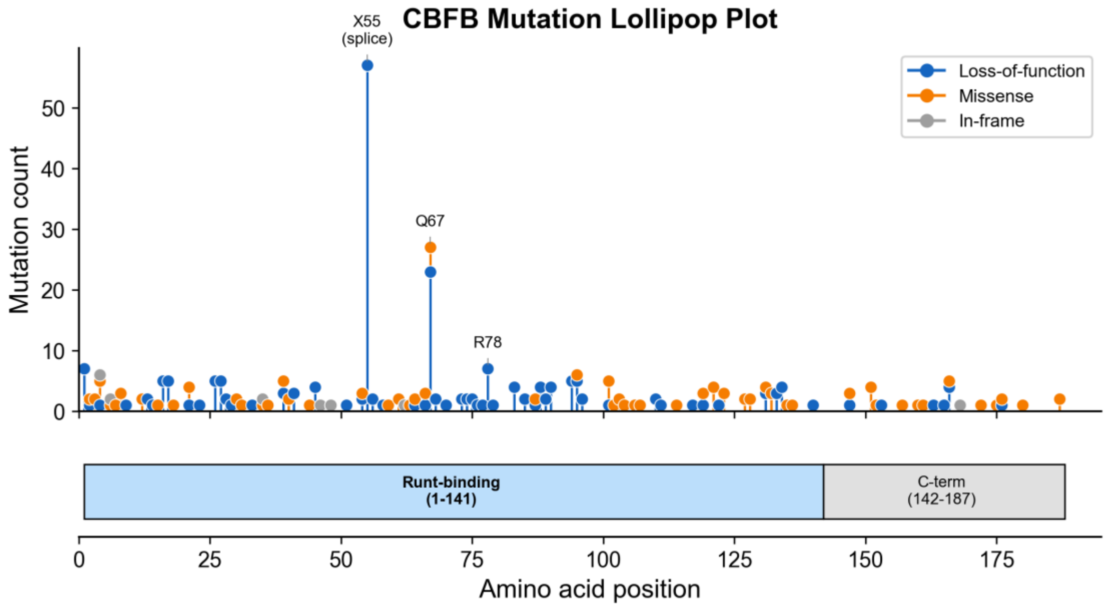


**Figure S9. CBFB lollipop plot.** Distribution of CBFB variants along the protein, color-coded by variant class. 91% of mutations map to the Runt-binding domain (amino acids 1–141), with a single hotspot at splice-site position 55 accounting for 18% of all mutations.

**Figure S10. Mutation-definition sensitivity forest.**

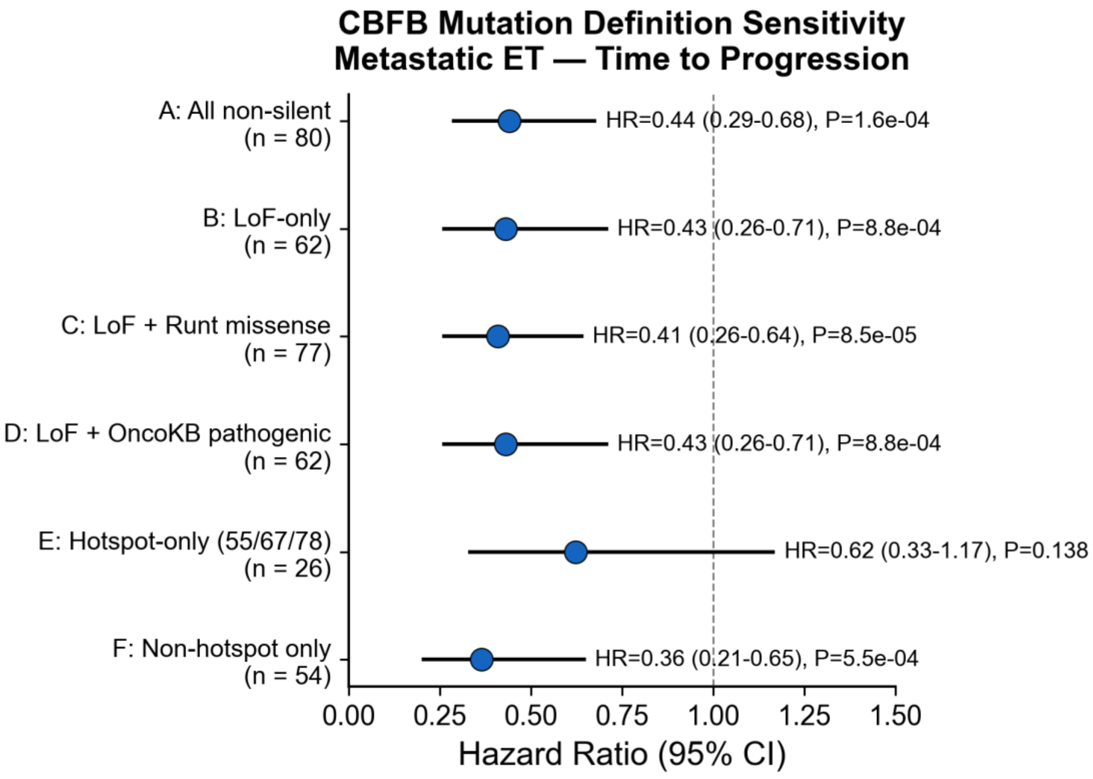

**Figure S10. Mutation-definition sensitivity forest.** CBFB hazard ratios across alternative mutation definitions: (A) all non-silent (primary); (B) loss-of-function only; (C) LoF + Runt-domain missense; (D) LoF + OncoKB-annotated pathogenic; (E) hotspot-only; (F) non-hotspot only. The narrow HR range (0.41–0.44) across the inclusive definitions confirms that the finding is driven by LoF biology.

**Figure S11. Structural mapping of CBFB and RUNX1 variants on the CBFβ–RUNX1 Runt domain–DNA ternary complex.**

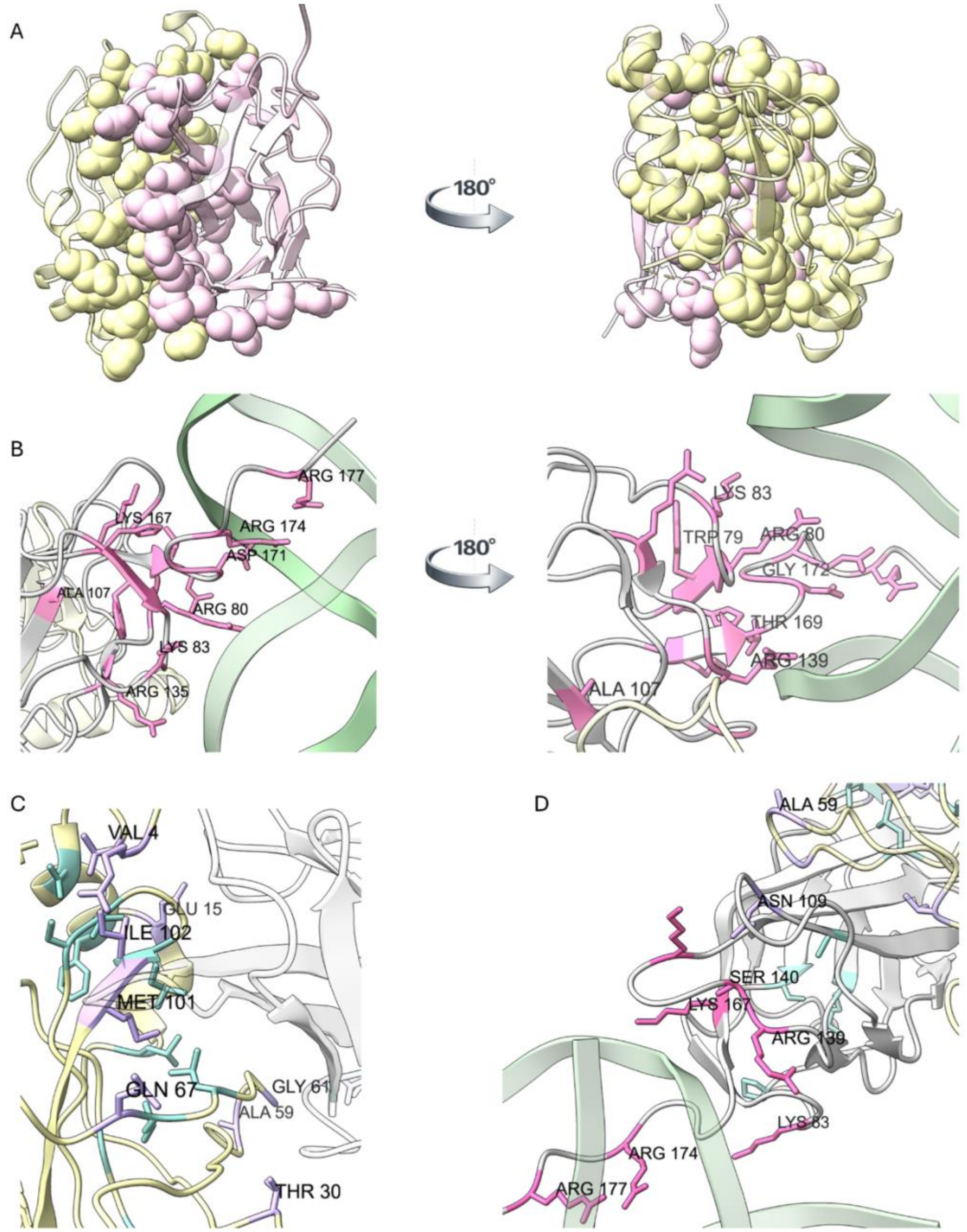

**Figure S11. Structural mapping of CBFB and RUNX1 variants on the CBFβ–RUNX1 Runt domain–DNA ternary complex.** (A) Interface residues (spheres) in the RUNX1 (yellow) CBFB (pink) complex (PDB 1H9D). (B) Germline pathogenic RUNX1 variants from ClinVar (pink, sticks) mapped onto RUNX1 (gray) DNA (green) and CBFB (yellow) interfaces (PDB 1IO4). (C-D) Somatic CBFB missense variants from MSK-CHORD mapped onto the CBFB-RUNX1 interfaces (purple, sticks), and RUNX1-DNA interface (pink, sticks), other variants shown in blue (PDB 1H9D).

### Supplementary Tables

##### Table ST1. Baseline characteristics by CBFB mutation status across the four cohorts.

| Cohort | Variable | CBFB mut | WT | P |
|---|---|---|---|---|
| Met ET | Age (median) | 58 | 60 | 0.89 |
| Met ET | N_met_sites (median) | 1 | 2 | $<10^{-15}$ |
| Met ET | Liver mets (%) | 6.2% | 17.7% | 0.01 |
| Met ET | Bone mets (%) | 25.0% | 45.5% | $6.0\times10^{-4}$ |
| Met ET | ILC (%) | 25.0% | 12.4% | 0.003 |
| Met ET | TP53 (%) | 7.5% | 25.3% | $5.0\times10^{-4}$ |
| Met ET | PIK3CA (%) | 52.5% | 40.7% | 0.05 |
| Adj ET | Age (median) | 69 | 65 | 0.14 |
| Adj ET | ILC (%) | 7.4% | 11.3% | 0.51 |
| Adj ET | TP53 (%) | 9.3% | 21.9% | 0.04 |
| Adj ET | PIK3CA (%) | 59.3% | 45.2% | 0.06 |

*P values from Wilcoxon rank-sum (continuous) or Fisher exact (categorical) tests. No clinically meaningful imbalances were observed between CBFB-mutated and CBFB-wild-type patients.*

##### Table ST2. Multivariable Cox models for CBFB across treatment cohorts.

| Model | Variable | N | HR | CI lower | CI upper | P |
|---|---|---|---|---|---|---|
| Met ET TTP — Age-adjusted | CBFB | 1197 | 0.44 | 0.29 | 0.67 | $1.6\times10^{-4}$ |
| Met ET TTP — Age-adjusted | CURRENT_AGE_DEID | 1197 | 1.00 | 0.99 | 1.01 | 0.89 |
| Met ET OS — Age-adjusted | CBFB | 1197 | 0.49 | 0.28 | 0.86 | 0.01 |
| Met ET OS — Age-adjusted | CURRENT_AGE_DEID | 1197 | 1.01 | 1.00 | 1.02 | 0.07 |
| Adj ET RFS — Age-adjusted | CBFB | 861 | 0.53 | 0.32 | 0.88 | 0.01 |
| Adj ET RFS — Age-adjusted | CURRENT_AGE_DEID | 861 | 0.98 | 0.98 | 0.99 | $6.9\times10^{-6}$ |
| Adj ET OS — Age-adjusted | CBFB | 861 | 0.52 | 0.28 | 0.97 | 0.04 |
| Adj ET OS — Age-adjusted | CURRENT_AGE_DEID | 861 | 0.98 | 0.97 | 0.99 | $9.8\times10^{-5}$ |
| Met ET TTP — Age+ILC-adjusted | CBFB | 1197 | 0.46 | 0.30 | 0.70 | $3.7\times10^{-4}$ |
| Met ET TTP — Age+ILC-adjusted | CURRENT_AGE_DEID | 1197 | 1.00 | 0.99 | 1.01 | 0.73 |
| Met ET TTP — Age+ILC-adjusted | IS_ILC | 1197 | 0.70 | 0.54 | 0.91 | 0.008 |
| Met ET OS — Age+ILC-adjusted | CBFB | 1197 | 0.49 | 0.28 | 0.85 | 0.01 |
| Met ET OS — Age+ILC-adjusted | CURRENT_AGE_DEID | 1197 | 1.01 | 1.00 | 1.01 | 0.08 |
| Met ET OS — Age+ILC-adjusted | IS_ILC | 1197 | 1.05 | 0.79 | 1.41 | 0.72 |
| Adj ET RFS — Age+ILC-adjusted | CBFB | 861 | 0.54 | 0.32 | 0.88 | 0.01 |
| Adj ET RFS — Age+ILC-adjusted | CURRENT_AGE_DEID | 861 | 0.98 | 0.98 | 0.99 | $5.3\times10^{-6}$ |
| Adj ET RFS — Age+ILC-adjusted | IS_ILC | 861 | 1.12 | 0.83 | 1.51 | 0.45 |

| Model | Variable | N | HR | CI lower | CI upper | P |
|---|---|---|---|---|---|---|
| Adj ET OS — Age+ILC-adjusted | CBFB | 861 | 0.52 | 0.28 | 0.98 | 0.04 |
| Adj ET OS — Age+ILC-adjusted | CURRENT_AGE_DEID | 861 | 0.98 | 0.97 | 0.99 | $9.5 \times 10^{-5}$ |
| Adj ET OS — Age+ILC-adjusted | IS_ILC | 861 | 1.07 | 0.73 | 1.55 | 0.74 |
| Met ET TTP — Full genomic | CBFB | 1197 | 0.50 | 0.33 | 0.77 | 0.002 |
| Met ET TTP — Full genomic | CURRENT_AGE_DEID | 1197 | 1.00 | 1.00 | 1.01 | 0.57 |
| Met ET TTP — Full genomic | IS_ILC | 1197 | 0.78 | 0.60 | 1.01 | 0.06 |
| Met ET TTP — Full genomic | TP53 | 1197 | 1.69 | 1.41 | 2.02 | $2.1 \times 10^{-8}$ |
| Met ET TTP — Full genomic | ESR1 | 1197 | 2.03 | 1.53 | 2.70 | $1.2 \times 10^{-6}$ |
| Met ET OS — Full genomic | CBFB | 1197 | 0.53 | 0.31 | 0.93 | 0.03 |
| Met ET OS — Full genomic | CURRENT_AGE_DEID | 1197 | 1.01 | 1.00 | 1.02 | 0.04 |
| Met ET OS — Full genomic | IS_ILC | 1197 | 1.28 | 0.95 | 1.72 | 0.11 |
| Met ET OS — Full genomic | TP53 | 1197 | 2.21 | 1.76 | 2.79 | $1.7 \times 10^{-11}$ |
| Met ET OS — Full genomic | ESR1 | 1197 | 3.59 | 2.59 | 4.98 | $1.8 \times 10^{-14}$ |

*Nested multivariable Cox proportional hazards models. Model A: CBFB + age. Model B: + ILC histology. Model C: + TP53 + ESR1. HR = hazard ratio for CBFB mutation; 95% CI = 95% confidence interval.*

#### Table ST3. CBF-complex (CBFB or RUNX1) biomarker and CBF × PIK3CA co-mutation results.

| Cohort | Endpoint | Analysis | Group | N | n group | HR | CI lower | CI upper | P |
|---|---|---|---|---|---|---|---|---|---|
| Met ET | TTP | CBF univariate | CBF_COMPLEX | 1197 | 122 | 0.48 | 0.34 | 0.67 | $2.0 \times 10^{-5}$ |
| Met ET | TTP | CBF×PIK3CA 4-group | CBF_only | 1197 | 60 | 0.73 | 0.49 | 1.10 | 0.13 |
| Met ET | TTP | CBF×PIK3CA 4-group | PIK3CA_only | 1197 | 435 | 0.99 | 0.84 | 1.18 | 0.95 |
| Met ET | TTP | CBF×PIK3CA 4-group | Both | 1197 | 62 | 0.27 | 0.15 | 0.49 | $1.7 \times 10^{-5}$ |
| Met ET | OS | CBF univariate | CBF_COMPLEX | 1197 | 122 | 0.60 | 0.40 | 0.91 | 0.02 |
| Met ET | OS | CBF×PIK3CA 4-group | CBF_only | 1197 | 60 | 0.78 | 0.45 | 1.34 | 0.37 |
| Met ET | OS | CBF×PIK3CA 4-group | PIK3CA_only | 1197 | 435 | 1.27 | 1.02 | 1.58 | 0.03 |
| Met ET | OS | CBF×PIK3CA 4-group | Both | 1197 | 62 | 0.57 | 0.31 | 1.04 | 0.07 |
| Met Chemo | TTP | CBF univariate | CBF_COMPLEX | 995 | 45 | 0.94 | 0.63 | 1.40 | 0.76 |
| Adj ET | RFS | CBF univariate | CBF_COMPLEX | 861 | 82 | 0.60 | 0.41 | 0.88 | 0.009 |
| Adj ET | RFS | CBF×PIK3CA 4-group | CBF_only | 861 | 35 | 0.88 | 0.53 | 1.48 | 0.634 |
| Adj ET | RFS | CBF×PIK3CA 4-group | PIK3CA_only | 861 | 350 | 0.96 | 0.79 | 1.15 | 0.642 |
| Adj ET | RFS | CBF×PIK3CA 4-group | Both | 861 | 47 | 0.45 | 0.26 | 0.78 | 0.005 |
| Adj ET | OS | CBF univariate | CBF_COMPLEX | 861 | 82 | 0.76 | 0.49 | 1.19 | 0.23 |

| Cohort | Endpoint | Analysis | Group | N | n group | HR | CI lower | CI upper | P |
|---|---|---|---|---|---|---|---|---|---|
| Adj ET | OS | CBF×PIK3CA 4-group | CBF_only | 861 | 35 | 1.33 | 0.77 | 2.30 | 0.31 |
| Adj ET | OS | CBF×PIK3CA 4-group | PIK3CA_only | 861 | 350 | 0.92 | 0.71 | 1.18 | 0.50 |
| Adj ET | OS | CBF×PIK3CA 4-group | Both | 861 | 47 | 0.39 | 0.18 | 0.82 | 0.01 |
| Adj Chemo | RFS | CBF univariate | CBF_COMPLEX | 566 | 51 | 0.83 | 0.58 | 1.19 | 0.302 |

*CBF-complex biomarker = patients with mutations in CBFB or RUNX1. Results presented for univariate Cox regression and four-way CBF × PIK3CA stratification.*
